# Common health conditions in childhood and adolescence, school absence, and educational attainment: Mendelian randomization study

**DOI:** 10.1101/19012906

**Authors:** Amanda Hughes, Kaitlin H Wade, Frances Rice, Matt Dickson, Alisha Davies, Neil M Davies, Laura D Howe

## Abstract

**Objectives:** To assess the causal relationship of different health conditions in childhood and adolescence with educational attainment and school absence.

**Design:** Longitudinal observational study and Mendelian randomization (MR) analyses.

**Setting:** Avon Longitudinal Study of Parents and Children (ALSPAC), a population sample of children from South-West England born in 1991-1992.

**Participants:** 6113 unrelated children with available GCSE records and genetic data (50% female).

**Exposures:** Six common health conditions with known genetic markers measured at age 10 (primary school) and 13 (mid-secondary school). These were: symptoms of Attention-Deficit Hyperactivity Disorder (ADHD), Autism Spectrum Disorder (ASD), depression, asthma, migraines and BMI. Genetic liability for these conditions and BMI was indexed by polygenic scores.

**Main outcome measures:** Educational attainment at age 16 (total GCSE and equivalents points score), school absence at age 14-16.

**Results:** In multivariate-adjusted observational models, all health conditions except asthma and migraines were associated with poorer educational attainment and greater school absence. Substantial mediation by school absence was seen for BMI (e.g. 35.6% for BMI at 13) and migraines (67% at age 10), with more modest mediation for behavioural and neurodevelopmental measures of health. In genetic models, a unit increase in genetically instrumented BMI z-score at age 10 predicted a 0.19 S.D. decrease (95% CI: −0.28 to −0.11) in attainment at 16, equivalent to around a 1/3 grade difference in each subject. It also predicted 8.6% more school absence (95% CI:1.3%, 16.5%). Similar associations were seen for BMI at age 13. Consistent with previous work, genetic liability for ADHD predicted lower educational attainment, but did not clearly increase school absence.

**Conclusions:** Triangulation across multiple approaches supported a causal, negative influence of higher BMI on educational attainment and school absence. Further research is required to understand the mechanisms linking higher BMI with school absence and attainment.

**What is already known on this topic:**

- On average, children with common health conditions have worse educational attainment
- It is unclear whether all health-attainment and health-absenteeism associations are causal, or reflect confounding by social and economic circumstances
- We do not know how much health-related school absenteeism contributes to these associations

**What this study adds:**

- Results support a negative influence of high BMI in secondary school on educational attainment (GCSEs) and absenteeism
- Absenteeism substantially mediated BMI-GCSE associations, suggesting a target for intervention
- There was less evidence for causal effects of Autism Spectrum Disorder, depressive symptoms, asthma or migraines on attainment and absenteeism
- Contribution of absenteeism to ADHD-GCSE associations was modest, suggesting interventions should target other mechanisms

## INTRODUCTION

Good health in childhood is positively associated with later socioeconomic outcomes including educational attainment (1-3), but the extent to which associations are causal is unclear. Health in childhood also shows consistent social patterning by family background, with worse health for less advantaged groups(4). This implies that health may play a key role in intergenerational transmission of socioeconomic (dis)advantage(5), suggesting an avenue for potential intervention(4). It also brings challenges for identification of causal mechanisms linking health to socioeconomic outcomes over the life-course(6), as socioeconomic circumstances in childhood may influence both attainment and health independently. This paper focuses on the impact of child and adolescent health on educational attainment: an aspect of socioeconomic position usually established in early adulthood, with extensive evidence of protective effects for later health(7).

Childhood and adolescent health encompasses behavioural, emotional and physical aspects, across which both the size of effect on educational outcomes and the pathways involved may differ(8). For example, Attention-Deficit Hyperactivity Disorder (ADHD) is related to lower educational attainment(9-11), with mixed evidence for depressive symptoms (12-17). There is marked variability in academic ability observed for children with Autism Spectum Disorder (ASD)(18, 19). Evidence is mixed for the effect on attainment of body weight(20-26), asthma(27-31) and rarer health conditions(27-29, 32, 33). School absence may play a role in the link between health and education but its contribution likely differs by condition(34-36). Migraine(30, 37) and depression appear linked to school absence(38), but evidence is mixed for asthma(27, 31, 35), ADHD (28, 39), ASD(28, 40) and obesity(41-44). Retrospective reporting of absence by children or parents(45) means estimates from previous studies may be affected by recall bias and measurement error(31).

Relationships of health, school absence and educational attainment are vulnerable to confounding. For instance, associations of obesity with social disadvantage(46) and poorer psychological health(47) may confound BMI-attainment associations(23), since disadvantage and poor psychological health may influence weight and attainment independently. Reverse causation may exaggerate associations with mental health(17). Approaches have been developed to circumvent these problems by using genetic variants (single nucleotide polymorphisms, or SNPs) associated with health conditions as proxies or instrumental variables. Since SNPs are assigned at conception, associations with SNPs cannot be due to reverse causation or classical kinds of confounding(48). Such studies support a causal influence of ADHD on educational attainment(9, 49), but report inconclusive effects for BMI(25, 26). Results for ASD have been null(49), or pointed to effect heterogeneity, with a positive relationship between ASD and attainment for high-functioning subgroups(50). Fewer studies have examined depression with null results reported(51). These genetic methods have not yet been applied to the influence of asthma or migraines on attainment.

To further understand these relationships, we applied a range of genetic methods to compare the impact of six aspects of childhood and adolescent health on educational attainment: ADHD, ASD, depressive symptoms, BMI, asthma, and migraines. Causality was explored in several ways. In an English birth cohort(52), we assessed associations of attainment with polygenic scores representing genetic liability for health conditions, and used polygenic scores as instrumental variables (one-sample Mendelian randomization). We explore mediation of associations by school absence and avoid the recall bias likely to have affected previous studies by using linked records for absence and educational attainment. We consider health in late primary school (approximately age 10) and mid-secondary school (approximately age 13), to examine whether influence of health on attainment varies with age. Finally, we conducted two-sample Mendelian randomization for the same conditions and educational attainment in independent adult samples.

## METHODS

### One and two-sample Mendelian Randomization

To overcome confounding and reverse causality, a range of approaches have been developed which use genetic variants (typically SNPs) associated with health. Multiple SNPs associated with a health condition can be combined into a polygenic risk score (PGS) representing genetic liability for a condition. Relative to single SNPs, this improves statistical power by explaining more variation in a health exposure of interest. In one-sample Mendelian randomization, an effect size for the causal influence of the exposure is estimated by using the PGS as an instrumental variable for the exposure in a two-stage least-squares model. Two-sample Mendelian randomization, in contrast, requires only summary-level results from genome-wide association studies(GWAS)(53). This compares associations of individual SNPs with an exposure (here, a health condition) on one hand and an outcome (here, educational attainment) on the other. If the exposure-outcome relationship is causal, the same SNPs should associate with both.

### Study participants

Individual-level data was obtained from the Avon Longitudinal Study of Parents and Children (ALSPAC), a birth cohort of children born in the south-west of England with estimated birth dates between April 1991 and December 1992. Inclusions and exclusions for this analysis are shown in Supplementary Figure 1. The total ALSPAC sample comprised 15,454 pregnancies, with 14,901 children alive at 12 months. Questionnaire and clinic data were obtained from mothers and children from pregnancy onwards, with detailed description given elsewhere(52, 54). After excluding related individuals, there were 7856 unrelated ALSPAC participants with genetic data, of whom 6113 had GCSE records. Multiple imputation with chained equations (m=50) was used to impute missing health measures (Supplementary Table 1), absences and covariates.

### Measures

#### Health in childhood and adolescence

ADHD symptoms were based on the hyperactivity subscale of the Strengths and Difficulties Questionnaire (SDQ-HI), as completed by mothers, when children were aged 9 and 13. The SDQ is a validated screening tool for psychiatric disorders in the relevant age group(55). Depressive symptoms were measured using the short-form Mood and Feelings Questionnaire (MFQ)(56) completed by children at ages 10 and 13. For autism, the parent-reported Social Communication Disorder Checklist (SCDC) at ages 10 and 13 was used to derive a continuous measure of autistic social traits (57). BMI (in kg/m^2^) was based on measured height and weight from clinic assessments at ages 10 and 13. From this, measures were standardized to the 1990 UK Growth Reference according to gender and age. Resulting z-scores, representing S.D. difference from the reference mean, were included in models as continuous variables. Presence of asthma in the past 12 months was defined using mother’s reports of diagnoses, medication use and wheezing symptoms, at ages 10 and 13. At age 10, mothers were asked if their children had experienced migraine, from which a binary indicator was derived. A later measure of migraine was not available. Please note that the study website contains details of all the data that is available through a fully searchable data dictionary and variable search tool: http://www.bristol.ac.uk/alspac/researchers/our-data/

#### Educational attainment, school absence, and covariates

Information on educational attainment and school absences came from linkage to the National Pupil Database (NPD). We consider educational outcomes at the end of year 11 (end of Key Stage 4), when most participants were aged 16 and which marked the end of compulsory education in the UK at the time. In all analyses, standard errors were clustered by school. We used the total GSCE and equivalents points score, based on a pupil’s best 8 GCSE or equivalent subjects. This is a continuous measure ranging from 0 to 540. One grade difference in one GCSE subject equates to 6 points, such that 5 grade Cs is worth 200 points and 8 A*s worth 464. A small number of scores above 464 reflect pupils who took AS levels early. More information is available from the Department of Education(58). School absence data was available for the academic years 2006-7 through 2008-9, corresponding to different school years for participants enrolled in ALSPAC (birth dates span 21 months). Records covered early September until the end of May. Information on absences was therefore available across the sample on absences in year 11, for the majority of participants on absences in year 10, but only for a minority in year 9 (Supplementary Table S1). We therefore considered school absence during the two years of key stage 4 (years 10 and 11), by imputing each separately and calculating an average post-imputation.

#### Polygenic scores

ALSPAC children were genotyped using the Illumina HumanHap550 platform, and standard quality control procedures applied. Individuals were excluded for gender mismatches, minimal or excessive heterozygosity, disproportionate individual missingness (>3%) and insufficient sample replication (IBD<0.8). Individuals with non-European ancestry were removed. SNPs with a minor allele frequency of <1%, call rate of < 95% or evidence of Hardy-Weinberg disequilibrium (p-value<5×10^−7^) were removed. Cryptic relatedness was measured as proportion of identity by descent (IBD>0.1). Imputation was performed using Impute v2.2.2 to the 1000Genomes reference panel, and SNPs with poor imputation quality (infoscore<0.08) removed.

GWAS were used to identify SNPs associated with ADHD, ASD, depression, asthma and migraine, and with BMI. We obtained SNP associations for ADHD(59), depression(51), ASD(50), asthma(60) and migraine(61) from GWAS including child and adult-onset conditions, since GWAS specifically of child-onset conditions were unavailable. For BMI, we used genetic variants associated with adult BMI from the most recent and largest BMI GWAS(62). A GWAS of BMI in children exists, but ALSPAC comprised a substantial component of the discovery sample(63), and such sample overlap can cause bias (64). When choosing SNPs to include in the polygenic scores, the conventional threshold of genome-wide significance of p<5×10^−8^ was applied, except for ASD. As too few SNPs meet that threshold to permit meaningful analysis, a more liberal threshold of p<5×10^−7^ was used. Within the set of SNPs which were available in ALSPAC and had passed standard quality control, we removed SNPs which were not independent (linkage disequilibrium clumping threshold r^2^=0.01, distance=10,000kb). Polygenic scores were calculated in PLINK 1.9 by summing each individual’s number of trait-increasing alleles. These were weighted by the regression coefficient for the allele’s association with the trait from the relevant GWAS – so that genetic variants with greater effects contributed more to the scores – and then standardized. Details of the GWAS and SNPs used are provided in Supplementary Figure 1, and Supplementary Tables S2 and S3.

### Statistical analysis

#### Analyses in ALSPAC

Analyses were conducted using STATAv15. The proportion of school sessions missed ranged from 0 to 0.79 (Table 1, Supplementary Table S4) with considerable skew, so for analysis was log-transformed after adding a constant of 0.01. Coefficients for absence are therefore expressed in terms of percentage change. For each aspect of health at age 10 and 13, linear regression was used to examine associations with GCSE points score and with logged school absence. All analyses were adjusted for gender and a number of potential confounders relating to family socioeconomic circumstances at birth: maternal age (in years) and parity, maternal education (from highest qualification, classified as none, CSE, vocational qualifications, O-level, A-level, or university degree), maternal smoking during pregnancy (yes/no), and maternal housing tenure (owner-occupier/council rented/private or housing association rented/other). Sensitivity analyses stratified by school type, restricting in turn to children attending mainstream state schools, independent (fee-paying) and other schools (community special schools, pupil referral units, further education colleges). Mediation analysis using STATA’s paramed package considered associations of health with GCSE points score via school absence (the indirect effect) and unexplained by school absence (the direct effect). Models were run separately within each imputed dataset and estimates combined across imputations.

**Table 1:** Descriptive Characteristics of Analytic Sample (N=6113)^a^.

|  | mean | SD | min | max |
| --- | --- | --- | --- | --- |
| Maternal age | 28.27 | 4.68 | 15.00 | 44.10 |
| SDQ hyperactivity score<sup>b</sup> at 10 (114 months) | 0.04 | 1.01 | -3.12 | 4.36 |
| SDQ hyperactivity score<sup>b</sup> at 13 (156 months) | 0.03 | 1.00 | -3.27 | 4.82 |
| MFQ score<sup>c</sup> at 10 (127 months) | 0.01 | 0.98 | -3.09 | 5.98 |
| MFQ score<sup>c</sup> at 13 (154 months) | 0.33 | 1.16 | -3.68 | 4.28 |
| SCDC scored at 10 (120 months) | 0.07 | 1.02 | -3.11 | 4.32 |
| SCDC scored at 13 (156 months) | 0.00 | 1.01 | -3.43 | 5.17 |
| BMI<sup>e</sup> z-score at 10 (127 months) | 0.05 | 1.02 | -3.21 | 5.98 |
| BMI<sup>e</sup> z-score at 13 (154 months) | 0.41 | 1.20 | -3.85 | 4.56 |
| GCSE capped points score | 332.34 | 87.36 | 0.00 | 540.00 |
| Percent of sessions absent, year 11 (age 15-16)<sup>f</sup> | 7.80 | 8.95 | 0.00 | 98.59 |
| Percent of sessions absent, year 10 (age 14-15)<sup>f</sup> | 7.10 | 8.28 | 0.00 | 90.39 |
| Percent of sessions absent, key stage 4 (age 14-16)<sup>f</sup> | 7.45 | 7.57 | 0.00 | 79.29 |
|  |  |  | % |  |
| gender | male |  | 49.99 |  |
|  | female |  | 50.01 |  |
| maternal educational qualifications | CSE or less |  | 16.81 |  |
|  | vocational |  | 9.49 |  |
|  | O level |  | 36.00 |  |
|  | A level |  | 24.29 |  |
|  | Degree |  | 13.41 |  |
| maternal housing tenure in pregnancy | mortgage/owned outright |  | 80.07 |  |
|  | council rented |  | 10.44 |  |
|  | private/other rented |  | 6.89 |  |
|  | other |  | 2.60 |  |
| maternal parity at child's birth | 0 |  | 44.90 |  |
|  | 1 |  | 36.16 |  |
|  | 2 |  | 14.00 |  |
|  | 3+ |  | 4.94 |  |
| mother smoked during pregnancy | no |  | 76.91 |  |
|  | yes |  | 23.09 |  |
| migraines at 10 | no |  | 95.16 |  |
|  | yes |  | 4.84 |  |
| asthma at 10 (128 months) | no |  | 87.90 |  |
|  | yes |  | 12.10 |  |
| asthma at 13 (157 months) | no |  | 88.05 |  |
|  | yes |  | 11.95 |  |
| school type<sup>g</sup> at key stage 4 (age 14-16) | mainstream state |  | 93.13 |  |
|  | independent |  | 5.45 |  |
|  | other |  | 1.42 |  |

<sup>a</sup>Analysis was restricted to unrelated ALSPAC participants with genetic data and linked GCSE records, N=6,113. Missing data in covariates, exposures and school absence was imputed using multiple imputation by chained equations. <sup>b</sup>SDQ=Strengths and Difficulties Questionnaire, for ADHD symptoms. <sup>c</sup>MFQ=Mood and Feelings Questionnaire, for depressive symptoms. <sup>d</sup>SCDC=Social Communication Disorder Checklist, for autistic social traits. <sup>e</sup>Using 1990 UK Growth Reference. Values represent standard deviation difference from age- and
gender-specific reference mean.<sup>f</sup> Absences were analysed as the number of half-day sessions recorded as missed, divided by the number of sessions on which data was available. For most participants, data was available each year for between 280 and 320 sessions, not the 390 of a standard school year, as records cover early September until the end of May. A small minority (2.3% in year 10, 4.8% in year 11) had data corresponding to fewer sessions.<sup>g</sup> Mainstream state schools: community, voluntary controlled or aided, foundation, city technology college, academy. Other schools: community special, pupil referral unit, further education college

Linear regression was used to examine associations between each of the polygenic scores with GCSE points score and with school absence. Genetic models were adjusted only for gender and 20 principal ancestry components. With a valid instrument, adjustment for confounders of observational associations is not necessary, and can introduce bias if those factors were not included in the GWAS used as the source of the gene variant-exposure associations(65). Where there was evidence of an association and the polygenic scores were shown to be sufficiently strong instruments (first-stage F-statistics >10), one-sample MR analyses were run using each of the polygenic scores as an instrument for the corresponding measured health condition at age 10 and 13. A concern in MR studies is pleiotropy, which can bias exposure-outcome causal estimates. This is when alleles related to the exposure (e.g. BMI) influence the outcome (e.g. GCSEs) via pathways other than through the exposure. Validity of instruments was checked using Stata’s MRRobust package. This applies two-sample MR methodology to the SNPs included in each PGS, producing MR-median, MR-modal and MR-Egger estimates(66).

#### Summary-level Mendelian Randomization

Using the TwoSampleMR package in R(67), summary-level MR analyses were performed to assess the causal effect of asthma, migraine and BMI on educational attainment. SNPs associated with educational attainment came from the most recent GWAS of years of schooling in European-ancestry individuals(68), except for BMI where an earlier education GWAS was used (69) to avoid sample overlap which could bias results. Details of the GWAS used MR are given in Supplementary Tables 1 and 2.

## RESULTS

### Socioeconomic background, GCSEs, and school absence

GCSEs scores and school absences varied by gender and by child’s socioeconomic background (Supplementary Table S5). Girls’ GCSE capped points scores were on average higher than boys: 345.6 (95% CI: 338.7, 352.5) compared to 319 (95%CI: 311.0, 327.1), but school absence was also slightly higher for girls: 7.8% (95%CI: 7.1%, 8.4%) compared to 7.1% (95% CI: 6.6%, 7.7%) for boys. Maternal education was positively associated with attainment, and negatively associated with school absence. Children whose mothers had a degree had an average GCSE point score of 393.2 (95% CI: 387.2, 399.1) and average school absence of 6.0% (95% CI: 4.6%, 7.4%). For children whose mothers had a Certificate of Secondary Education or no qualifications, average GCSE points score was 275.6 (95% CI:266.9, 284.4) and average school absence 9.8% (95% CI: 9.0%,10.7%). As expected, GCSE points scores were negatively associated with school absence. For example, adjusted for gender, an increase in absence corresponding to an extra day/year at key stage 4 was associated with −2.7 (−3.3,−2.0) fewer GCSE points.

### Phenotypic models: health, GCSEs and school absence

In phenotypic models (Figure 1, Table 2), all aspects of child and adolescent health were associated with GCSE points score except for migraines and asthma. All were associated with school absence (Figure 2, Table 3). Depressive symptoms at age 10 showed a considerably stronger association with GCSEs than depressive symptoms at 13 (GCSE points scores: −14.75; 95% CI: −17.45, −12.06 compared to −5.39; 95% CI: −7.93, −2.85 per SD MFQ score). Otherwise, associations did not differ substantially by age. In mediation analyses, associations between all aspects of health and educational attainment were mediated by school absence expect for asthma, where indirect and total effects went in opposite directions at age 10, and indirect and direct effects in opposite directions at 13 (Figure 3, Supplementary Table S6). Percent of associations mediated by absence varied, with the lowest for ADHD (6.1% at age 10, 7.9% at 13) and the highest for BMI (48.5% at age 10, 35.7% at 13) and migraine (67.4% at age 10). (Figure 3, Supplementary Table S4). Results were similar restricting to participants in mainstream state schools. For other school types, small numbers of participants led to imprecise estimates (Supplementary Tables S7 and S8).

**Figure 1:**
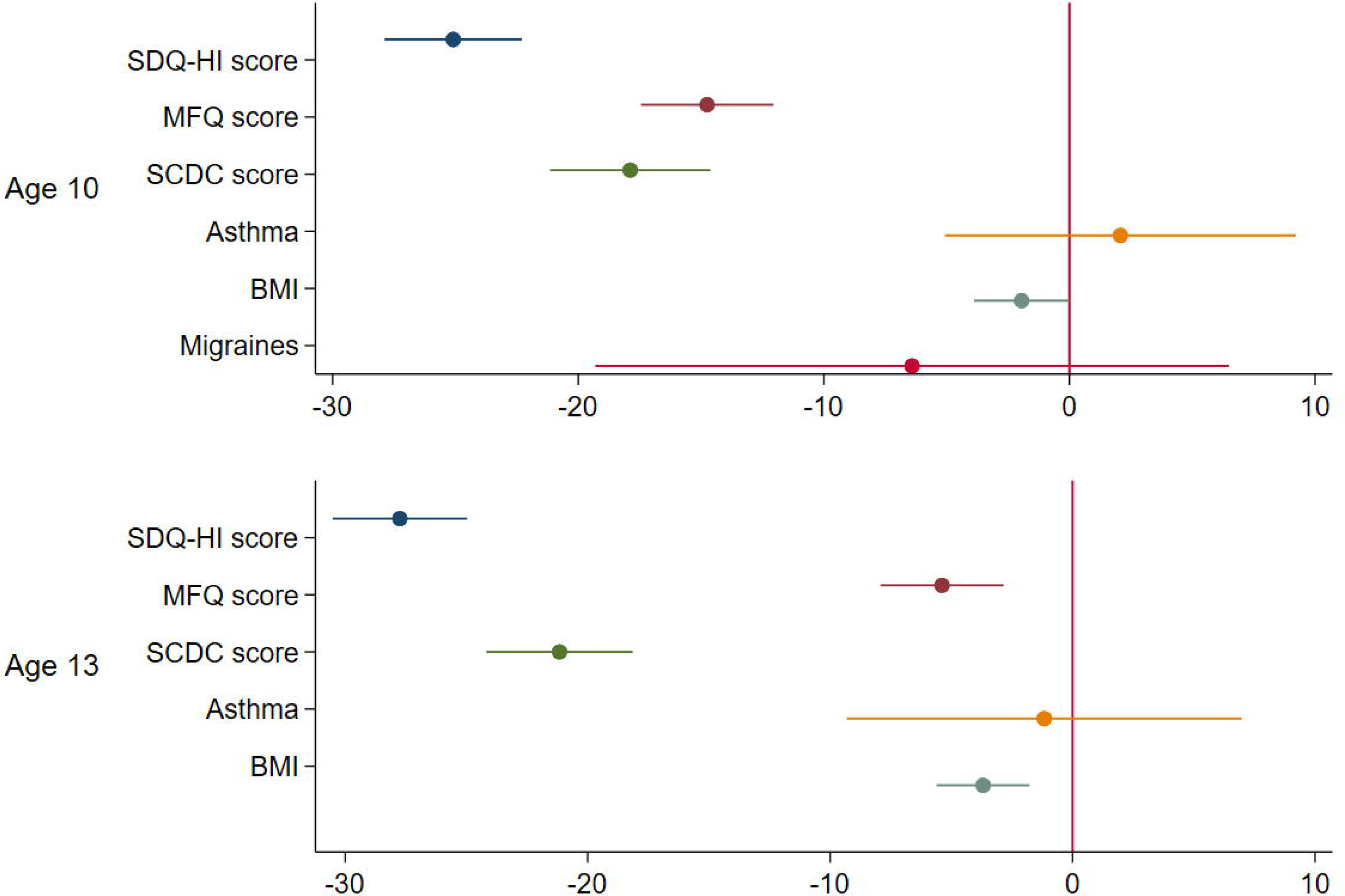
Phenotypic associations of childhood health and educational attainment at age 16. N=6113. Educational attainment: GCSE capped points score, range 0-540, mean 332.3. Coefficients represent change in GCSE points with presence of the health condition, or per S.D. increase in continuous exposures. SDQ-HI: Strengths and Difficulties Questionnaire hyperactivity subscale, for ADHD symptoms; MFQ: Mood and Feelings Questionnaire, for depressive symptoms; SCDC: Social Communication Disorder Checklist, for autistic social traits. BMI z-score: based on 1990 UK Growth Reference, values represent S.D. difference from age- and gender-specific reference mean

**Table 2:** Phenotypic associations: health in childhood and GCSE points score (range 0-540, mean=332.3, SD=87.4)^a^.

|  | Health at age 10 |  | Health at age 13 |  |
| --- | --- | --- | --- | --- |
|  | Beta | CI | Beta | CI |
| Standardized values of SDQ-HI score<sup>b</sup> | -25.09 | -27.89,-22.29 | -27.75 | -30.52,-24.97 |
| Standardized values of MFQ score<sup>c</sup> | -14.75 | -17.45,-12.06 | -5.39 | -7.93,-2.85 |
| Standardized values of SCDC score<sup>d</sup> | -17.88 | -21.15,-14.62 | -21.16 | -24.18,-18.15 |
| Migraines at age 10 | -6.41 | -19.31,6.49 |  |  |
| Asthma in past 12 months | 2.08 | -5.07,9.22 | -1.17 | -9.32,6.97 |
| BMI z-score<sup>e</sup> | -1.95 | -3.88,-0.01 | -3.70 | -5.61,-1.78 |

<sup>a</sup>N=6113. Coefficients represent change in GCSE capped points score adjusted for gender, maternal education, maternal housing tenure, maternal age, maternal parity, whether smoked in pregnancy. <sup>b</sup>SDQ-HI: Strengths and Difficulties Questionnaire hyperactivity subscale. <sup>c</sup>Mood and Feelings Questionnaire. <sup>d</sup>Social Communication Disorders Checklist. <sup>e</sup>Based on 1990 UK Growth Reference, values represent S.D. difference from age- and gender-specific reference mean

**Table 3:**
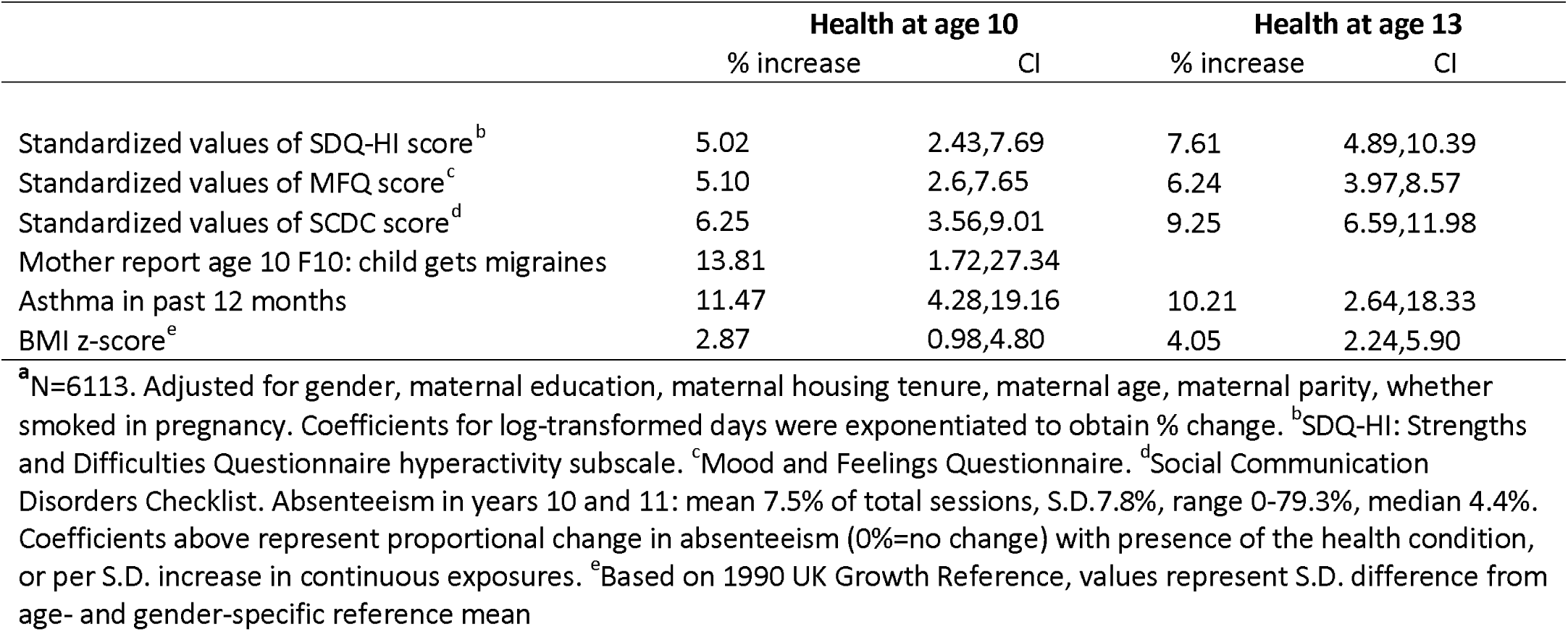
Phenotypic associations: health at 10 and 13 to absenteeism at age 14-16^a^.

**Figure 2:**
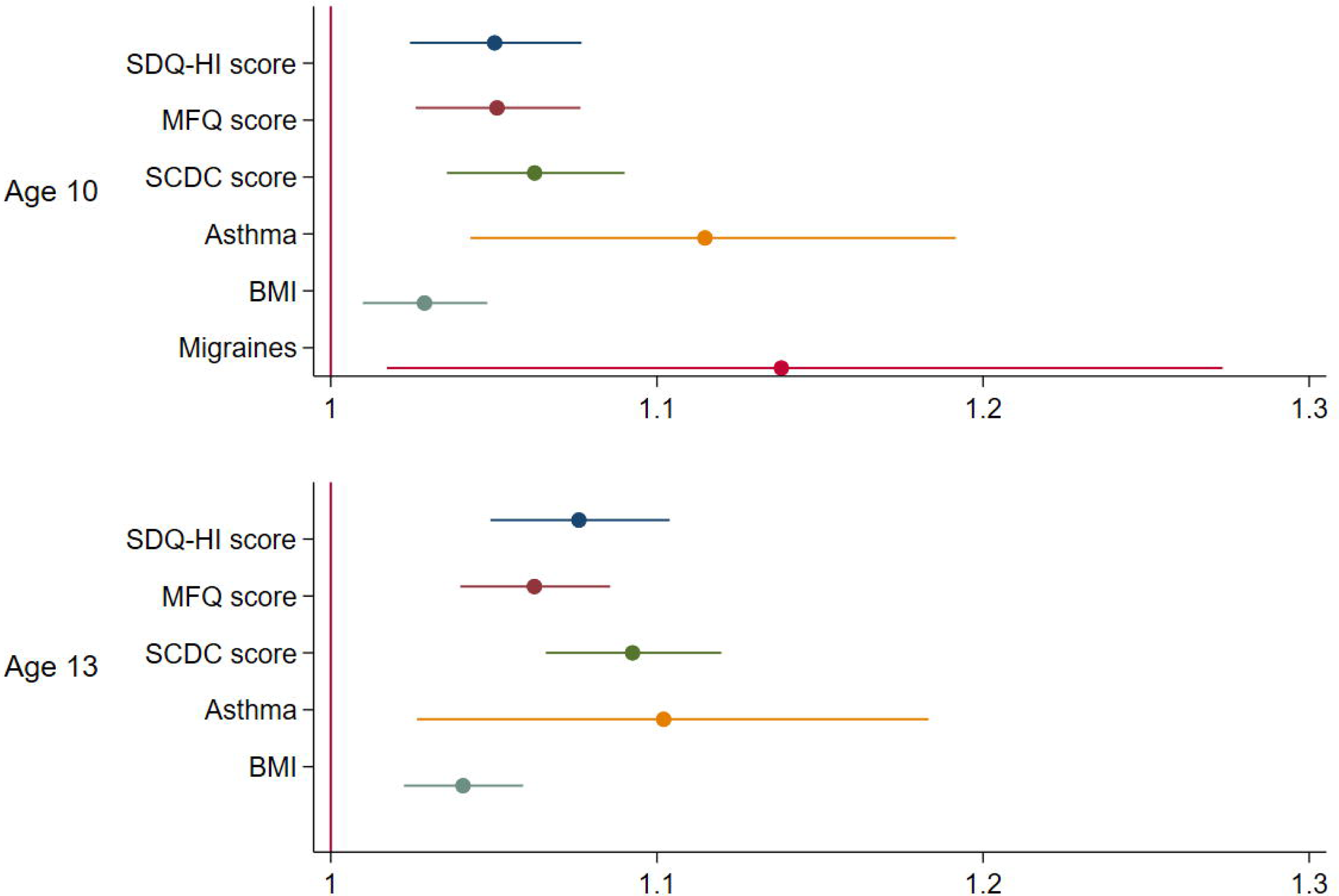
Phenotypic associations of childhood health and school absence at age 14-16. N=6113. Coefficients represent proportional change (1=no change) in year 10 and 11 absenteeism with presence of the health condition, or per S.D. increase in continuous exposures. Absenteeism at age 14-16: mean 7.5% of total sessions, S.D.7.8%, range 0-79.3%, median 4.4% SDQ-HI: Strengths and Difficulties Questionnaire hyperactivity subscale, for ADHD symptoms; MFQ: Mood and Feelings Questionnaire, for depressive symptoms; SCDC: Social Communication Disorder Checklist, for autistic social traits. BMI z-score: based on 1990 UK Growth Reference, values represent S.D. difference from age- and gender-specific reference mean

**Figure 3:**
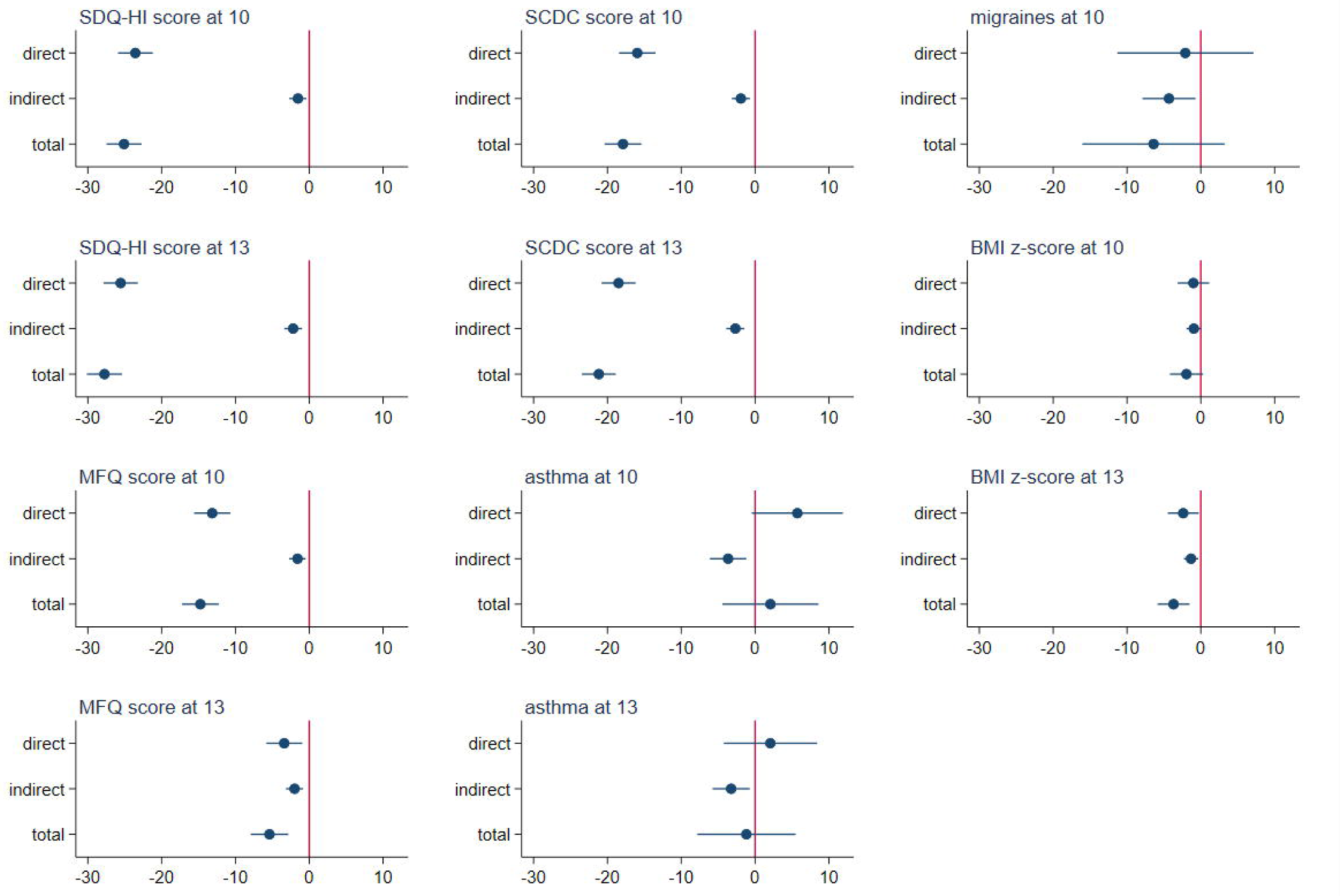
Mediation of Phenotypic associations of health with GCSEs by absenteeism at age 14-16. N=6113. GCSE points: mean 332.3, S.D. 87.4, range 0-540. Absenteeism at age 14-16: mean 7.5%, S.D.7.8%, range 0-79.3%. Coefficients represent change in GCSE points with presence of health condition, or per S.D. increase in continuous exposures. SDQ-HI: Strengths and Difficulties Questionnaire hyperactivity subscale, for ADHD symptoms; MFQ: Mood and Feelings Questionnaire, for depressive symptoms; SCDC: Social Communication Disorder Checklist, for autistic social traits. BMI z-scores: based on 1990 UK Growth Reference, values represent S.D. difference from age-and gender-specific reference means. Percent of total effects mediated by absenteeism: SDQ-HI at 10: 6.1%, SDQ-HI at 13: 7.9%, MFQ at 10: 10.9%, MFQ at 13: SCDC at 10: 10.8%, SCDC at 13: 12.6%, Migraines at 10: 67.4%, Asthma at 10: −175.8%, Asthma at 13: 277.1%, BMI z-score at 10: 48.5% 37.7%, BMI z-score at 13: 35.7%

### Genetic models: health, GCSEs and school absence

Predictive power of the polygenic scores varied considerably by health condition. The proportion of variance explained by the scores (R^2^ or pseudo-R^2^ for binary exposures) was 7.4% and 7.7% for BMI at 10-11 and 13, respectively, but less than 1% for migraine and asthma, and less than 0.1% for ADHD, ASD and depressive symptoms (Supplementary table S9). Tests for instrument strength (Supplementary Table S7) confirmed the BMI PGS could be used to instrument BMI z-scores, but the other PGSs could not be used as instruments.

For ADHD and BMI, higher values of the polygenic scores predicted lower GCSE points (Table 4, Figure 4). Specifically, each SD increase in the genetic liability for ADHD corresponded to a decrease of 2.70 (95% CI: −4.83, −0.58) GCSE points. A one SD increase in the BMI genetic score corresponded to a decrease of 5.37 (95% CI: −7.78, −2.96) GCSE points and a 2.70% (95% CI: 0.58%,4.86%) increase in school absence. Using the polygenic score for BMI as an instrumental variable showed that, for each unit increase in BMI z-score at age 10, GCSE points scores were 17.01 lower (95% CI: −24.76, −9.28) and absences were 8.60% greater (95% CI:1.27%, 16.47%). For each unit increase in age- and gender-standardized BMI at age 13, GCSE points scores were 16.12 lower (95% CI: −23.43, −8.81) and absences were 8.13% greater (95% CI: 1.22%, 15.52%).

**Table 4:** Association of polygenic scores with GCSEs and absenteeism at age 14-16^a^.

|  | GCSE<br>points | CI | Absences:<br>% increase | CI |
| --- | --- | --- | --- | --- |
| Standardized values of ADHD PGS | -2.70 | -4.83, -0.58 | -0.20 | -2.13,1.78 |
| Standardized values of depression PGS | 0.75 | -1.75, 3.25 | -0.80 | -2.84,1.29 |
| Standardized values of ASD PGS | -1.78 | -3.84, 0.28 | -0.55 | -2.60,1.54 |
| Standardized values of migraine PGS | -0.93 | -3.04, 1.18 | 1.36 | -0.630,3.39 |
| Standardized values of asthma PGS | -0.66 | -2.80, 1.48 | 1.15 | -0.66,3.00 |
| Standardized values of BMI PGS | -5.37 | -7.78, -2.96 | 2.70 | 0.58,4.86 |

<sup>a</sup>N=6113. Adjusted for gender and PC1-PC20. GCSE points score: range 0-540, mean 332.3, S.D.87.4. Coefficients for absences represent proportional change in absenteeism (0%=no change) per S.D. increase in the polygenic score.

**Figure 4:**
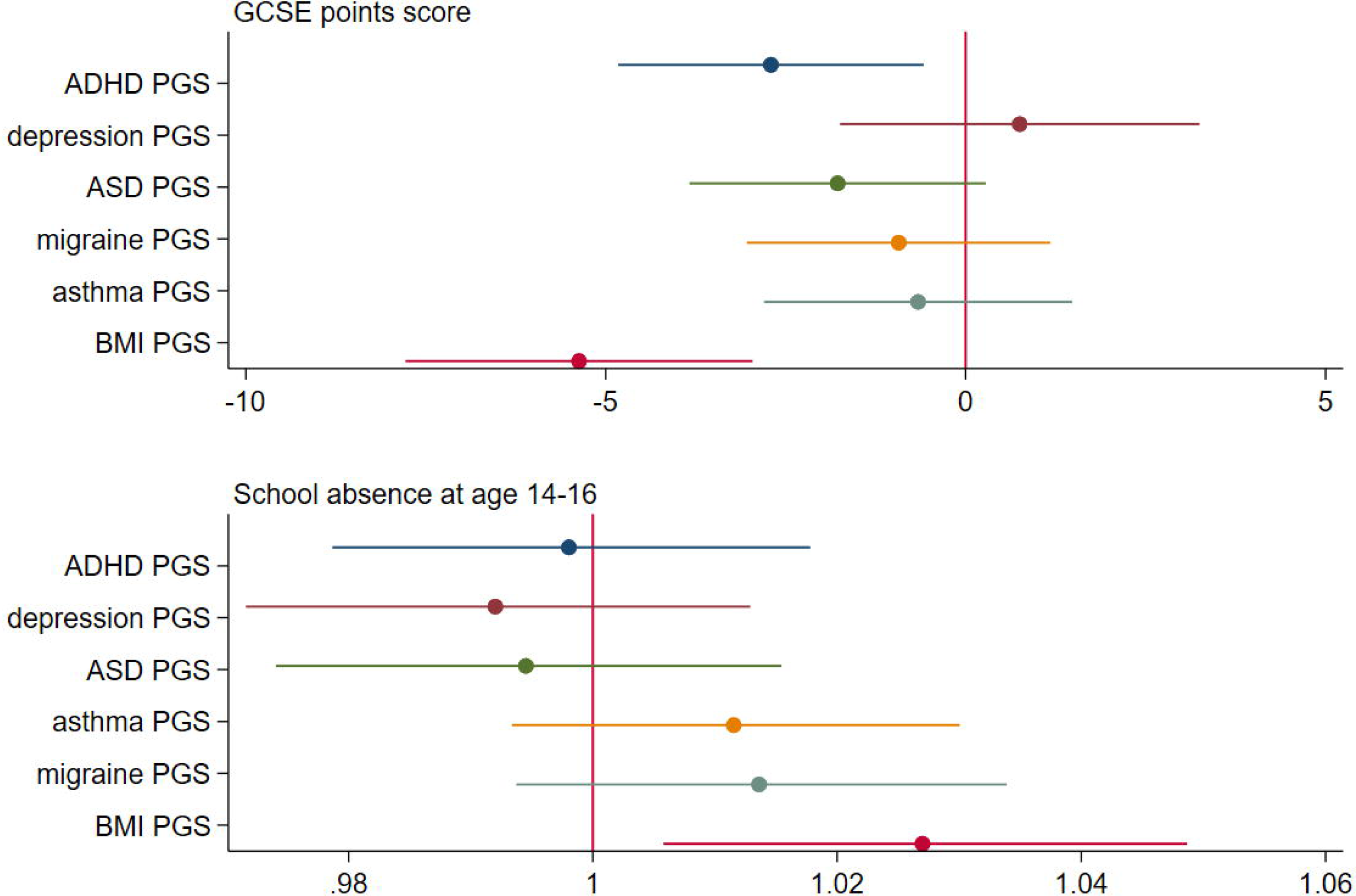
Association of polygenic scores with GCSEs and absenteeism at age 14-16. N=6113. Polygenic scores are standardized. GCSE points score: mean 332.3, S.D. 87.4, range 0-540. Absenteeism in years 10 and 11: mean 7.5% of total sessions, S.D.7.8%, range 0-79.3%, median 4.4%. Coefficients represent proportional increase in school absence per standardized unit increase in PGS (1=no change)

**Figure 5:**
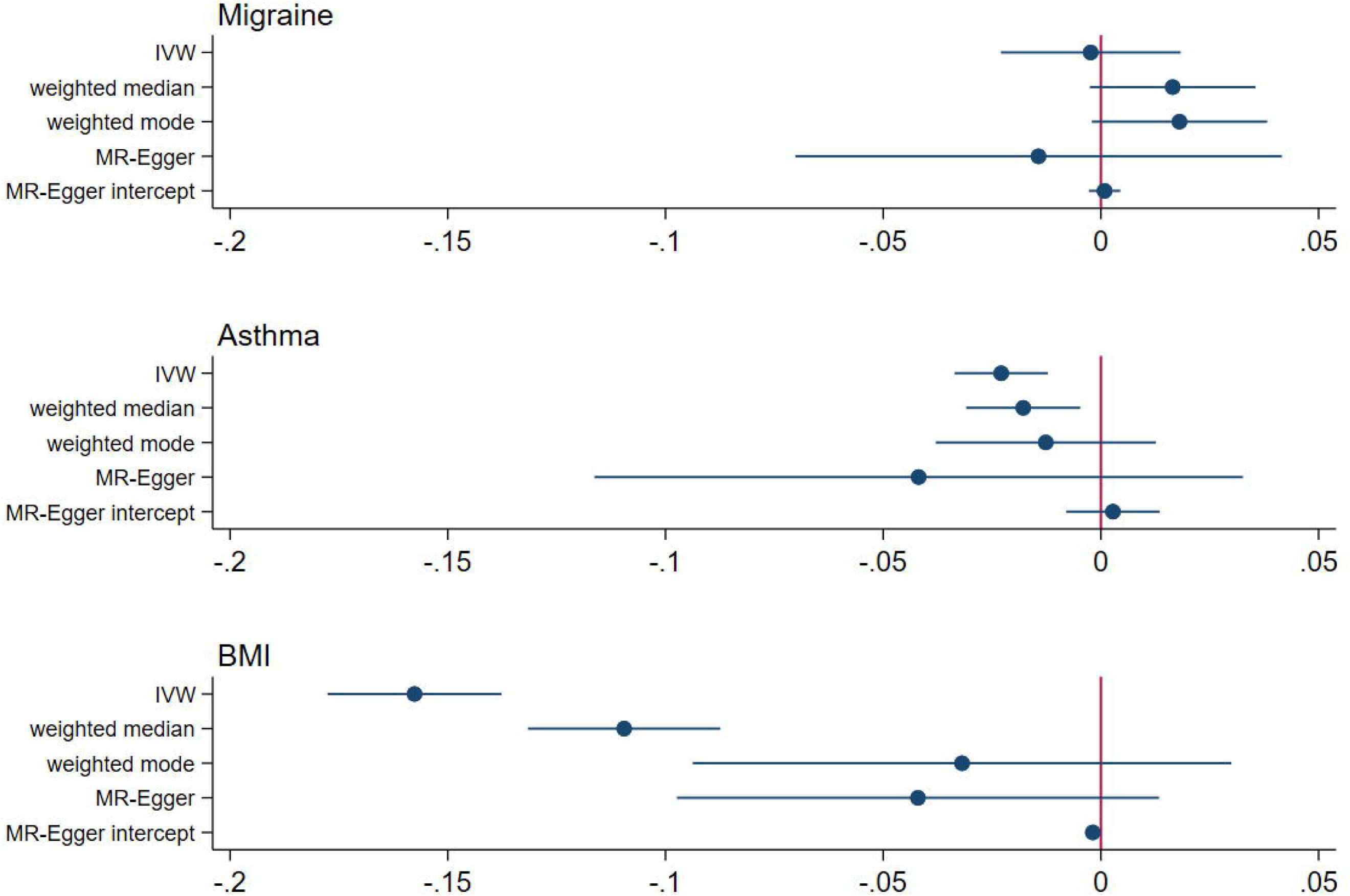
Results of Two Sample Mendelian Randomization. Associations with health and with educational attainment were taken from GWAS listed in Table S1. A non-zero value for the MR-Egger intercept indicates pleiotropy (influence of SNPs on the outcome of interest not via the exposure).

Results were similar restricting to participants in mainstream state schools, while small numbers of participants again led to imprecise estimates for other school types (Supplementary Table S10). Tests of instrument validity applying two-sample methodology in ALSPAC (Supplementary Table S11) were consistent with main results, and there was no evidence of bias due to pleiotropy for associations with GCSEs. There was evidence of pleiotropy for effect of BMI on absenteeism (MR-Egger intercept 0.004, p=0.02), but additional SNP-specific checks(70) could not identify particular SNPs responsible (Supplementary Table S12). Results did not differ using a BMI PGS excluding 24 SNPs identified as outliers in Two-Sample MR (Supplementary Table S13).

Results of two-sample MR analyses using published GWAS for educational attainment (years of schooling) were broadly consistent with results from ALSPAC (Table 5). Previous two-sample MR analysis reported evidence of an effect of ADHD(49) but not ASD(49) or depression(51). Our analyses supported an influence of BMI, with a one-unit increase in BMI associated with 0.16 (p<0.001) and 0.11 (p<0.001) fewer years of schooling in IVW and weighted median models respectively. These models also support a negative influence of asthma on years of schooling (−0.02, p<0.001 and −0.02, p=0.01 respectively). There was little evidence of a causal impact of migraine. For BMI, the MR-Egger constant (−0.002, p<0.001) indicated an influence of pleiotropy. Outliers were therefore identified by comparing SNP-specific estimates with the overall IVW estimate, and analyses repeated with these SNPs excluded. This did not change conclusions (supplementary table S14). We also checked associations of these SNPs with other phenotypes but none stood out as clear confounders (supplementary table S14).

**Table 5:**
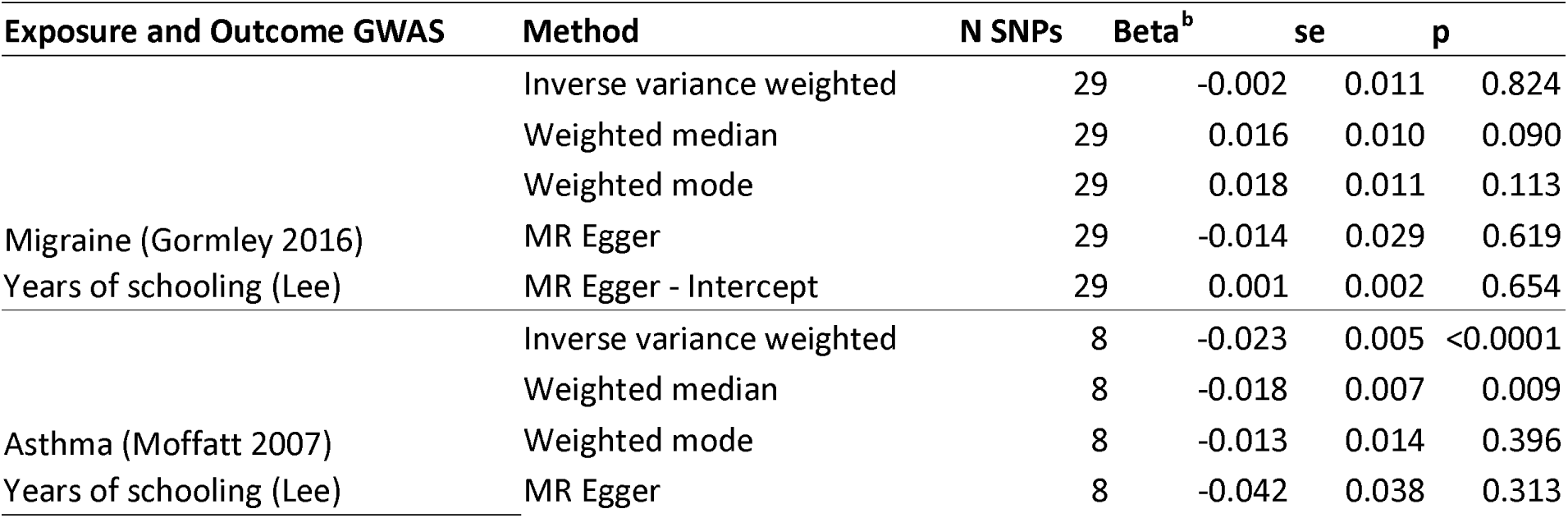
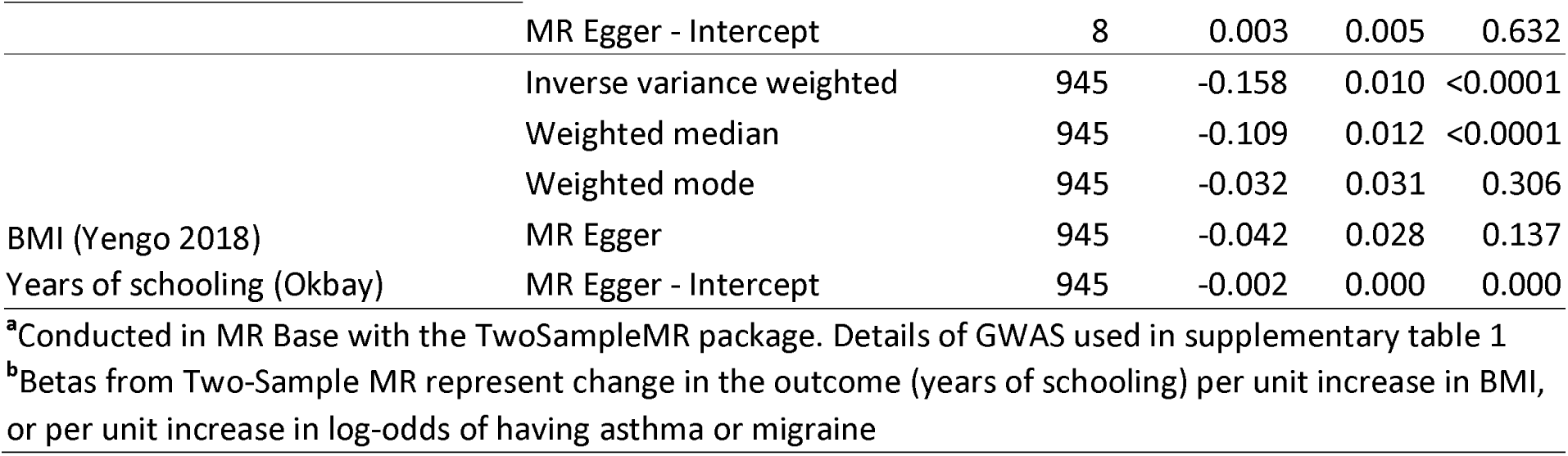
Results from Two-Sample Summary-level Mendelian Randomization^a^.

## DISCUSSION

In phenotypic analysis, all aspects of adverse health at ages 10 and 13 predicted greater school absences between age 14 and 16, and all aspects of adverse health except asthma and migraines predicted lower educational attainment at 16. Lack of associations of asthma and migraine with GCSEs may reflect binary measures unable to capture the full range of symptoms, or misclassification of these exposures as diagnosis can occur later. However, these associations are vulnerable to reverse causation and residual confounding by family- and individual-level characteristics not captured by covariates. In line with previous work in ALSPAC(9), polygenic scores representing genetic liability for ADHD were associated with worse GCSEs, but they were not associated with greater school absence. Polygenic scores for higher BMI were associated with both. Models using the PGS to instrument BMI supported these findings, and results shown to be robust to exclusion of outlier SNPs identified from Two-Sample MR. MR analyses using ALSPAC found some evidence of pleiotropy in BMI-absenteeism associations. However, we could not identify individually pleiotropic SNPs. SNP-specific estimates were imprecise, and mechanisms should be explored in samples large enough to investigate SNP-specific pathways more fully. Results from two-sample summary-level Mendelian randomization also supports a negative causal impact of higher BMI on educational attainment, despite the exposure and outcome being measured in individuals who were considerably older than ALSPAC participants. Results indicate that, for BMI, associations with attainment and absenteeism do not reflect only confounding. That IV coefficients were larger in magnitude than OLS coefficients for the (negative) influence of BMI on GCSEs may point to an offsetting mechanism causing suppression of effects in observational models. In UK children, both thinness and obesity are associated with deprivation(71), and a nonlinear relationship between BMI and socioeconomic hardship could lead to underestimation of the impact of high BMI on educational attainment. IV estimates could also be inflated due to family-level processes, such as the influence of parents’ genotype on offspring via environmental pathways(72) which may bias MR estimates based on samples of unrelated individuals(73).

A key strength of this study is triangulation across several methodological approaches to investigate if associations are causal. Negative associations of depressive symptoms at age 12(74), ADHD symptoms in preschool(75) and obesity at 11 and 16 with GCSEs have been previously demonstrated in the ALSPAC cohort(21). Our results support a causal interpretation of the latter two. They are consistent with studies into educational impact of ADHD and depression using within-sibling comparison (10, 12), which addresses confounding at the level of the family, but not confounding at the level of the individual, such as by comorbidities. Two-sample MR suggested an influence of asthma, as well as ADHD and BMI, on educational attainment. Although not apparent in ALSPAC, this was observed in a recent study using UK Biobank participants, where genetically-instrumented asthma corresponded to a 17% lower probability (CI:-25.3% to −8.7%) of holding a degree(76). Since in the two-sample analysis educational attainment was measured in a substantially older population(68), the discrepancy could reflect a cohort effect, with better treatment available to younger cohorts, or asthma diagnoses made in adulthood.

A limitation concerns the relative strength of the genetic instruments, which was considerably greater for BMI than other aspects of health. For ADHD, ASD and depression, the low proportion of variance in the phenotype explained by the polygenic scores limited the degree to which genetic methods could be meaningfully applied. For ASD, a more lenient threshold was required for SNPs included in the PGS. Thus, results of genetic models for ASD and depressive symptoms should not be interpreted as evidence of no effect. Rather, they indicate that associations may become clearer as the genetics of these conditions becomes better understood. Migraine could only be examined at age 10, but not 13, where effects may be greater. Recent evidence points to bias due to family-level processes even in genetic studies of BMI(73). It is possible that such effects have influenced our results, potentially overestimating BMI’s causal influence on attainment. Work in study populations with genetic data on related family members will be required to investigate this further. ALSPAC is not a national survey, and over-representation of more affluent groups and an under-representation of non-White minority ethnic groups(52) may limit generalizability of findings. A major strength was use of linked records for educational attainment and school absence, which meant associations were not influenced by recall bias. Results of mediation analysis are consistent with a recent UK study(45) reporting substantial mediation by parents’ reports of long-term absence (>1month) for any long-term health condition and a measure of mental health. Our findings indicate such results do not only reflect recall bias, and add to current knowledge by also showing mediation by absence for high BMI. A limitation is that information on absence was restricted to age 14-16, and absence earlier in secondary school may also affect attainment. Mechanisms other than absenteeism linking health to attainment are likely to be complex. For BMI, research has pointed to negative neurocognitive correlates of obesity, but this largely cross-sectional literature has not established a causal influence(77) and the evidence from longitudinal studies is much less clear(78). Socially-mediated processes by which weight could influence educational outcomes involve weight bias by teachers(79) and the impact of bullying by peers(80). Further work using both genetic and qualitative approaches will be required to unpick these mechanisms.

## Conclusion

In a UK-based cohort born in the early 1990s, our analyses supported a negative, causal influence of high BMI on educational attainment and school absence. Mediation analysis supported substantial mediation by absenteeism for BMI, and a smaller role for absenteeism in ADHD. The impact of health on attainment was not fully explained, highlighting the need for a better understanding of the social and biological mechanisms involved.

## Data Availability

The data concerns individual human subjects and therefore is not freely available, but can be obtained by researchers by applying to ALSPAC.

## Acknowledgements

We are extremely grateful to all the families who took part in this study, the midwives for their help in recruiting them, and the whole ALSPAC team, which includes interviewers, computer and laboratory technicians, clerical workers, research scientists, volunteers, managers, receptionists and nurses. The UK Medical Research Council and Wellcome (Grant ref: 102215/2/13/2) and the University of Bristol provide core support for ALSPAC. This publication is the work of the authors, who will serve as guarantors for the contents of this paper. A comprehensive list of grants funding is available on the ALSPAC website (http://www.bristol.ac.uk/alspac/external/documents/grant-acknowledgements.pdf). GWAS data was generated by Sample Logistics and Genotyping Facilities at Wellcome Sanger Institute and LabCorp (Laboratory Corporation of America) using support from 23andMe.

This research was specifically funded by the Health Foundation. The Medical Research Council (MRC) and the University of Bristol support the MRC Integrative Epidemiology Unit [MC_UU_12013/1, MC_UU_12013/9, MC_UU_00011/1]. The Economics and Social Research Council (ESRC) support NMD via a Future Research Leaders grant [ES/N000757/1]. LDH is supported by a Career Development Award from the UK Medical Research Council (MR/M020894/1). KHW is supported by the Elizabeth Blackwell Institute for Health Research, University of Bristol and the Wellcome Trust Institutional Strategic Support Fund [204813/Z/16/Z]. This work is part of a project entitled ‘Social and economic consequences of health: causal inference methods and longitudinal, intergenerational data’, which is part of the Health Foundation’s Efficiency Research Programme. The Health Foundation is an independent charity committed to bringing about better health and health care for people in the UK. No funding body has influenced data collection, analysis or its interpretation. This publication is the work of the authors, who serve as the guarantors for the contents of this paper.

Ethical approval for the study was obtained from the ALSPAC Ethics and Law Committee and the Local Research Ethics Committees. Consent for biological samples has been collected in accordance with the Human Tissue Act (2004). Informed consent for the use of data collected via questionnaires and clinics was obtained from participants following the recommendations of the ALSPAC Ethics and Law Committee at the time.

## Patient and public involvement

Since 2006 ALSPAC has an advisory panel of 30+ study participants who meet six times per year to provide insights and advice on study design, methodology and acceptability for participants. No participants were asked to advise on interpretation or writing up of results for this analysis. There are no specific plans to disseminate the results of the research to study participants, but the key findings from projects are made available on the ALSPAC website. Further information can be found at http://www.bristol.ac.uk/alspac/about/

## Author contributions

AH reviewed existing literature, prepared the data and carried out analysis. LH and NMD obtained funding for this work. All authors contributed to study design, interpreted results and revised the manuscript. AH is the guarantor for this work.

## Transparency declaration

The lead author affirms that this manuscript is an honest, accurate, and transparent account of the study being reported. No important aspects of the study, or discrepancies from work originally planned, have been omitted.

